# An Extended Susceptible-Exposed-Infected-Recovered (SEIR) Model with Vaccination for Predicting the COVID-19 Pandemic in Sri Lanka

**DOI:** 10.1101/2021.06.17.21258837

**Authors:** R. M. Nayani Umesha Rajapaksha, Millawage Supun Dilara Wijesinghe, Toms K. Thomas, Sujith P. Jayasooriya, B. M. W. Indika Gunawardana, W. M. Prasad Chathuranga Weerasinghe, Shalini Bhakta, Yibeltal Assefa

**Author notes:** **Corresponding Author** Dr. R. M. N. U. Rajapaksha, TP: /.

## Abstract

The role of modelling in predicting the spread of an epidemic is important for health planning and policies. This study aimed to apply a compartmental model for predicting the variations of epidemiological parameters in Sri Lanka. We used a dynamic Susceptible-Exposed-Infected-Recovered-Vaccinated (SEIRV) model and simulated potential vaccine strategies under a range of epidemic conditions. The predictions were based on different vaccination coverages (5% to 90%), vaccination-rates (1%, 2%, 5%) and vaccine-efficacies (40%, 60%, 80%) under different R_0_ (2,4,6). We estimated the duration, exposed, and infected populations. When the R_0_ was increased, the days of reduction of susceptibility and the days to reach the peak of the infection were reduced gradually. At least 45% vaccine coverage is required for reducing the infected population to mitigate a disastrous situation in Sri Lanka. The results revealed that when R_0_ is increased in the SEIRV model along with the increase of vaccination efficacy and vaccination rate, the population to be vaccinated is reducing. Thus, the vaccination offers greater benefits to the local population by reducing the time to reach the peak, exposed and infected population through flattening the curves.

## Introduction

### Background

#### COVID-19 situation

After 15 months from the Public Health Emergency of International Concern (PHEIC) declaration by the World Health Organization (WHO), Novel Corona Virus is still spreading throughout the world despite various degrees of movement restrictions and availability of multiple safe and effective vaccines [1]. According to the Weekly epidemiological update on COVID-19 on 11^th^ May 2021, there is a 4% reduction of the number of new COVID-19 cases and deaths globally, with over 5.5 million cases and over 90,000 deaths for the past week. Although there was a 13% reduction of the newly reported weekly caseload in the Eastern Mediterranean and a 23% reduction in Europe regions, South-East-Asia (SEA) shows upward trends for the 9^th^ consecutive week, which reported a further 6% increase as of 9^th^ May 2021. New deaths are increasing in SEA (15%) and Africa (3%), while India remains the primary concern as the contributing country for half of the global cases and nearly one-third of global deaths [2]. Similar to the regional situation, the number of cases is rising in Sri Lanka, with 2375 per day (14^th^ to 19^th^ May 2021). A total of 147,720 individuals are confirmed as infected by May 19, 2021[3,4]. Strain B.1.1.7 is a more transmissible variant that was initially detected in the United Kingdom, is currently circulating in Sri Lanka [5]. Therefore, early understanding of the epidemic and demand dynamics is fundamental in health planning and policymaking, especially when the resources are limited.

#### Compartmental models for prediction

Compartmental models can be used to project scenarios with various disease control measures individually or as a useful combination for evidence-based policy formulation and alteration. With the purpose of forecasting, different forecasting models are proposed by various academics and research groups. However, these forecasting models have their strengths and limitations. Therefore, they need to be interpreted cautiously. Furthermore, the underlying data are changing rapidly [6,7]. There are broad categories of mathematical models for COVID-19 forecasting’s, such as mass action compartmental models, structured metapopulation models and Agent-Based Network Models. Epidemiologists have been using mass action compartmental models over a period of hundred years which are famous for simplicity in both analysis and outcome assessment [8]. Importantly, from early December 2020, mass vaccination programs were started against the COVID-19 pandemic. Pfizer/BioNTech, Oxford/AstraZeneca, Janssen, Moderna and Sinopharm vaccines are included in the WHO Emergency Use Listing (EUL) and are widely used globally [9]. By 19^th^ May 2021, 9.35% of the world population has received at least one dose of the COVID-19 vaccine and 1.56 billion vaccine doses are already administered globally [10,11]. In Sri Lanka, 1,397,999 individuals (2.6% of the population) were vaccinated by at least one dose of vaccine out of Covishield, Sinopharm or Sputnik-V vaccines through mass vaccination programmes [3,12]. More importantly, public health interventions successfully limited the initial rapid transmission of COVID-19 in many countries. However, the epidemic recrudescence could be occurred due to the relaxation of these measures without achieving elimination or high levels of herd immunity risks [13].

#### SEIR model and extensions

Several compartmental models belong to the basic Susceptible-Infectious-Recovered (SIR) class. In the SIR model, the total population (N) is divided into compartments of Susceptible (S), Infectious (I) and Recovered (R). The SIR models are extended by adding an Exposed (E) compartment based on the same principle. It is assumed that every individual in the population is going through those four roles from susceptibility to recovery [Susceptible (S)-> Exposed (E)-> Infected (I)-> Recovered (R)]. Although real-life situations have some limitations, this has been used as a basic model for different epidemics [8,14]. Due to the proven effect of prevention of infection by vaccination, Vaccination (V) was also included in the SEIR model (a derivative of the classic SIR model), and the Susceptible-Exposed-Infected-Recovered-Vaccinated (SEIRV) forecasting model is formulated.

#### Vaccination rate, coverage, efficacy, and effectiveness

The vaccination rate against COVID-19 is one of the infection rate’s main determinants (along with the vaccine efficacy and effectiveness). It should be considered with the other factors related to the spreading of the disease, such as adherence to preventive measures. It was shown that the reducing infection rate begins with the different vaccination rates in different countries. It has been further observed that the infection rate after vaccination shows two trends: the inverted U-shaped trend and the L-shaped trend [15]. Sri Lanka started Covishield (AstraZeneca) as the COVID-19 vaccine for the country’s vaccination programme from 29^th^ January 2021 [16]. Even though the efficacy data of the different COVID-19 vaccines vary in different global studies, there should be a minimum of 50% efficacy for the WHO emergency use licensing of a COVID-19 vaccine [17]. Furthermore, the efficacy of the same COVID-19 vaccine can vary with the variant of the virus that depends on the vaccine schedule, such as whether only the first dose was taken, or the person has completed the schedule [18]. It was found that the effectiveness of the Covishield (AstraZeneca) vaccine after the first dose was 76% (95% CI: 61-85%) and the 86% (95% CI: 53-96%) following the completion of both doses against the alpha variant (B1.1.7) which is currently predominant variant in Sri Lanka [19].

### Aim of the study

The role of modelling in predicting the spread of an epidemic is important. Therefore, the present study aimed to construct a compartmental epidemiological model incorporating vaccination coverage, rate of the vaccination campaign, vaccine efficacies and applied a computational tool for predicting the evolution of different epidemiological variables for COVID-19 in Sri Lanka. We applied a dynamic SEIRV model for this purpose. The predictions were based on the SEIRV model without vaccination, evolution of infectious proportion under different vaccination coverages (5% to 90%), vaccination rates (1%, 2%, 5%) and vaccine efficacies (40%, 60%, 80%) at different R_0_ (2,4,6). The model was used to estimate the duration and infected population following different vaccination coverages.

## Method

### Compartmental epidemiological model

We constructed a compartmental epidemiological model (Fig 1) with vital dynamics describing the number of individuals in a fixed population who are susceptible to infection (S), exposed (E), infected (I), recovered (R), and vaccinated (V) (8,14). This simple deterministic model has several structural assumptions, including homogenous mixing of a closed population, no stratification of transmissibility by subpopulations, and complete and permanent immunity after natural infection.

**Fig 1.**
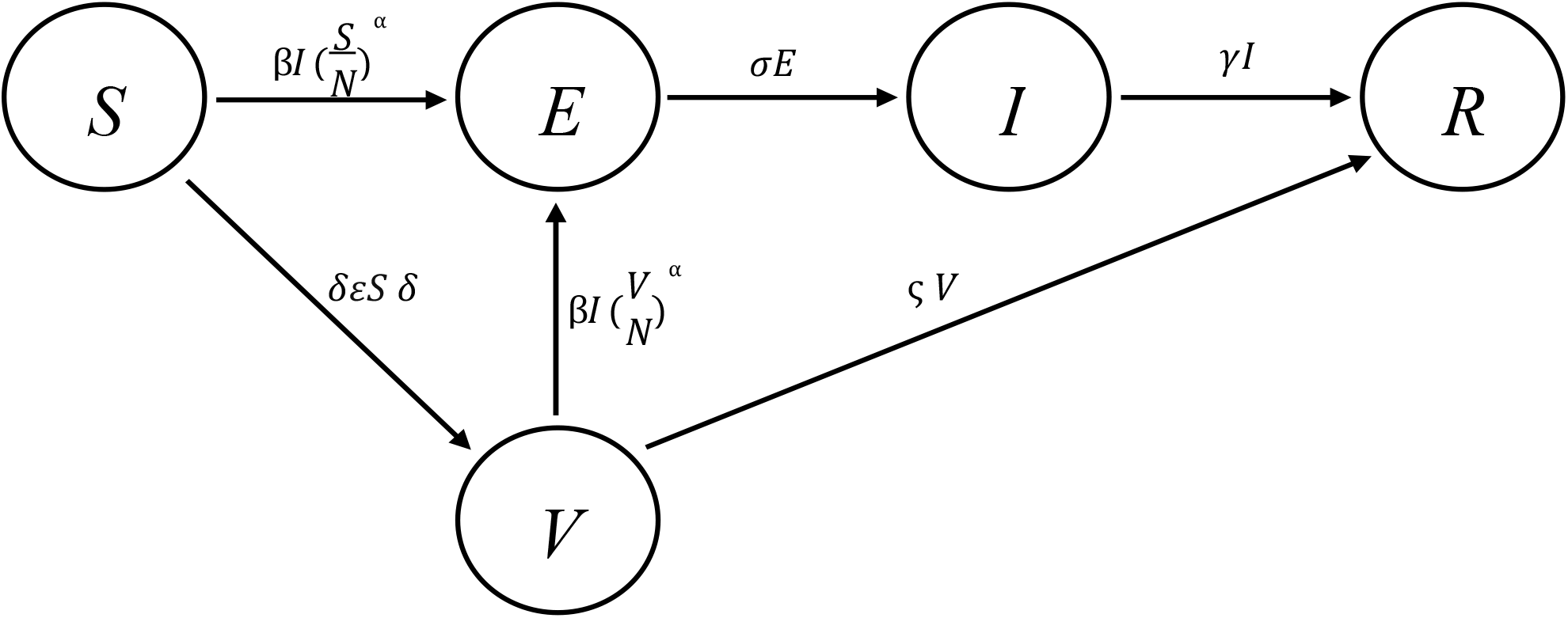
The SEIRV model with transition forces.

A set of ordinary differential equations governs the flow of individuals between compartments. We extracted publicly available data with permission from the official website of the Health Promotion Bureau of the Ministry of Health, Sri Lanka. The data was cross-checked with the daily update on the website of the Epidemiology Unit, Ministry of Health, Sri Lanka [3,4]. We used anonymized data for this analysis and extracted data relevant to cases reported from 11^th^ March 2020 to 15^th^ May 2021. Based on reported cases and the documented parameters, the model was validated for its ability to predict the number of cases for a period of 14 days in February 2021 and model fitting is illustrated in Supporting file I (S1 File).

Any personally identifiable data was not included in the analysis of this study. Python programming language was used for the development of the model. We have considered three hypothetical values for R_0_. Predictions for the SEIRV model were made when the R_o_ value is 2, 4 and 6 (Fig 1). Parameter beta (β) represents the transmission rate of the COVID-19. As one of the main assumptions, we assumed that the transmission rate β for both the susceptible and vaccinated populations is equal during the model development stage.

### Model equations

The flow of individuals through the compartments of the model is determined by a set of Ordinary Differential Equations (ODE). The time-variant parameters such as rate of change of susceptible population, exposed population, infected population, recovered population and vaccinated population is obtained by derivates with the following equations. Given the SEIRV model with transition forces, the ODEs are derived as below.

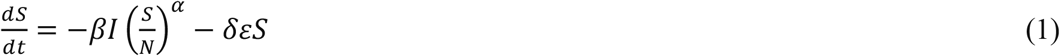

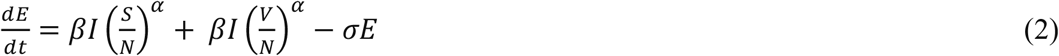

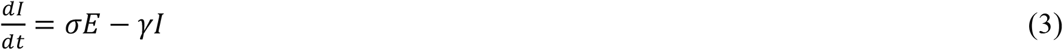

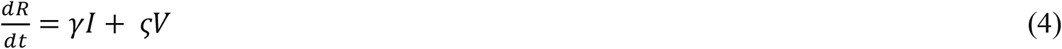

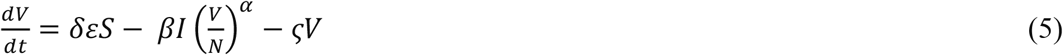

### Disease characteristics

The available COVID-19 data was used as the disease characteristics in this exploration (Table 1). However, there is still substantial uncertainty around these estimates and how they apply to a given setting, and there are insufficient data on which to base credible parameter distributions.

**Table 1.** Disease characteristics.

| Parameter | Modelled value | Reference |
| --- | --- | --- |
| Basic reproductive number (to predict) | 2, 4, 6 | Assumed |
| The latent period prior to infectivity | 3.2 days | [20] |
| Duration of infectivity | 8.5 days | [1, 9] |
| Time from infection to death | 22 days | [21] |
| Infection Fatality Ratio (IFR) | 0.66% | [2, 21] |

### Mass vaccination programme

We modelled an indiscriminate mass vaccination programme where 13,151 individuals received their first dose of vaccine against COVID-19 during the first 100 days of the vaccination campaign in Sri Lanka (as would be expected in the absence of a reliable and scalable method of determining either pre-existing immunity or active or incubating infection) [3]. The model parameters are presented in Table 2.

**Table 2.**
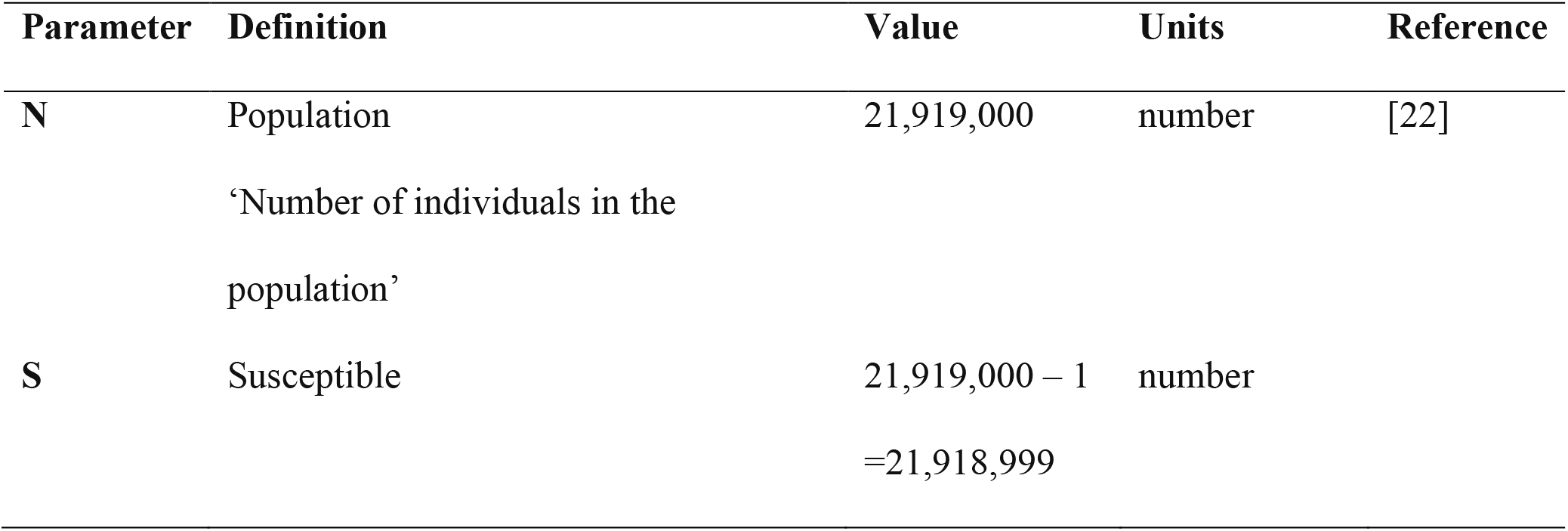
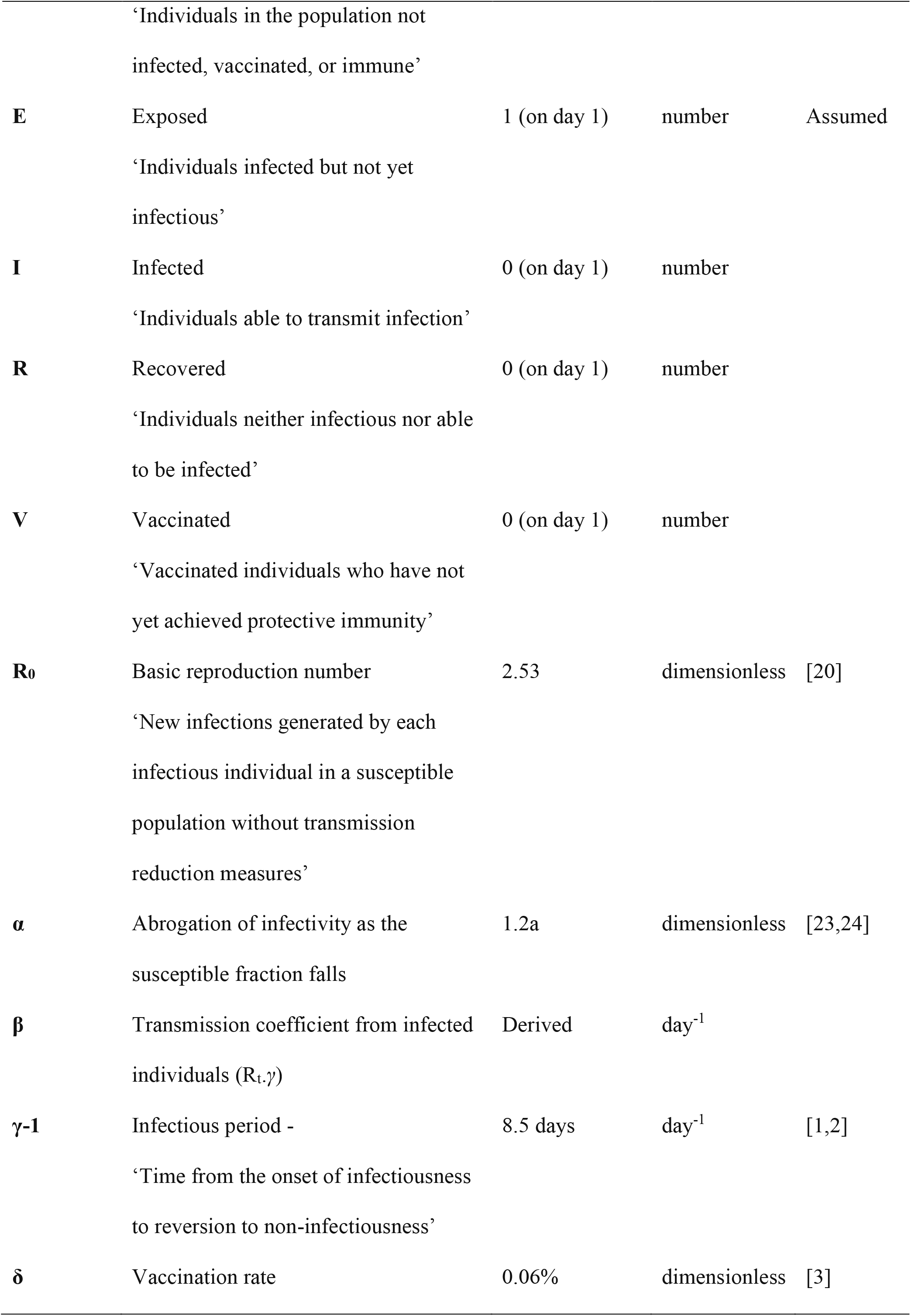
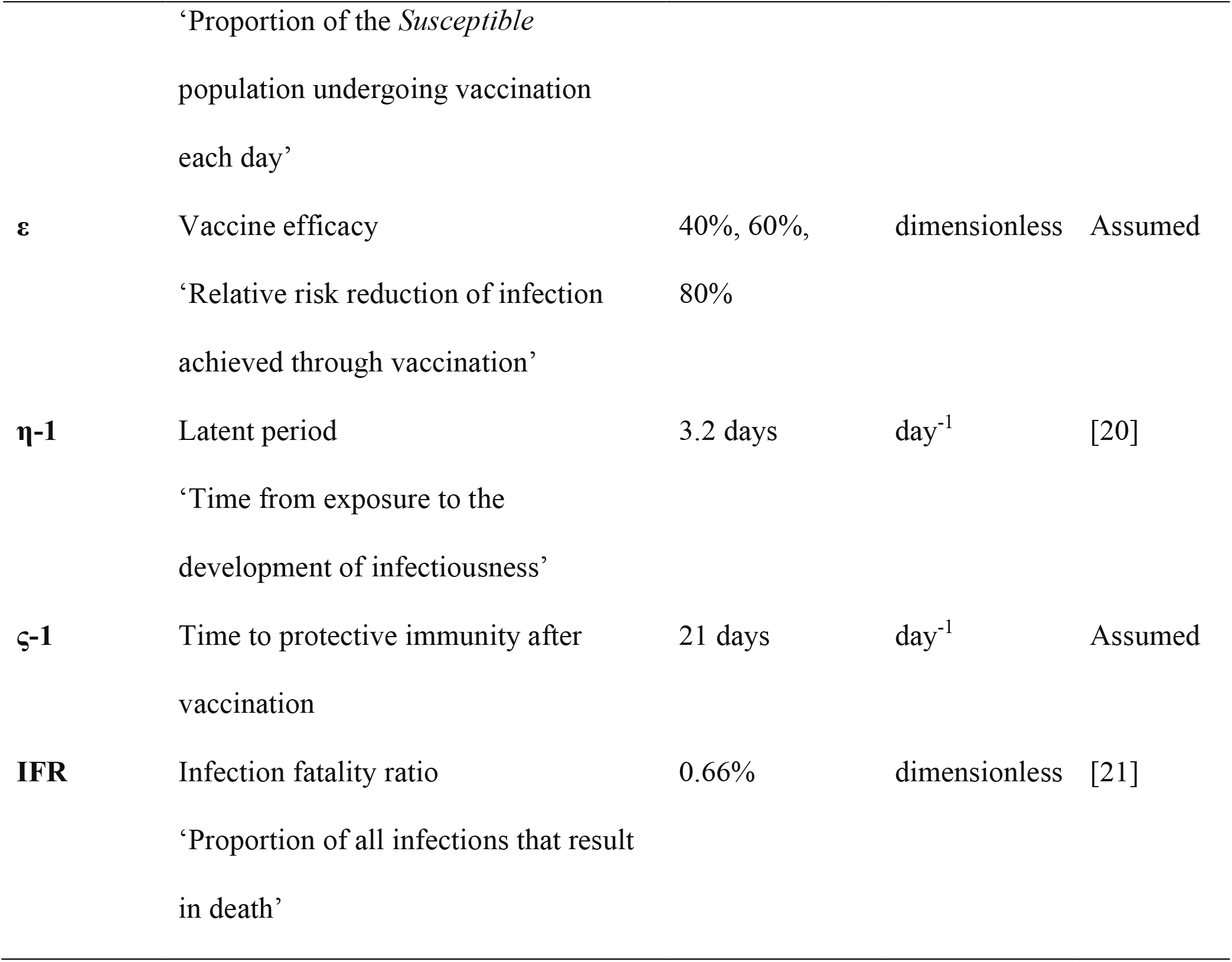
Values for the model parameters corresponding to the Sri Lankan COVID-19 situation.

### Vaccination coefficient in the model

A combination of ‘vaccination efficacy’ and ‘vaccination rate’ is known as ‘vaccination coefficient’ in the model. Notably, the vaccination efficacy and vaccination rate are determined by the health authorities. When the R_0_ and rate of vaccination are constant and vaccination efficacy is increased (40%, 60% and 80%), the vaccination coefficient will be increased in line with the increased efficacy. Therefore, it implies that the increase of vaccination coefficient would increase the vaccinated population while reducing the exposed and infectious population to reach its maximum within a smaller number of days. Moreover, when the R_0_ is constant, vaccination efficacy and rate of vaccination are increased, the vaccination coefficient is also increased. Therefore, the number of vaccinated populations from the susceptible population is also increased. As a result, the number of exposed, infectious populations declines and reaches the equilibrium level in the early days in the graphs, which indicates that the COVID-19 crisis can be controlled if we increase the efficacy and rate of vaccination. Under a lower level of vaccination coefficient, the equilibrium comes in later days of the graphs while it increases the exposed and infectious populations.

## Results

As in a pandemic like COVID-19, a compartmental model is a good approach for comprehension and analyzing epidemiological data. However, the model needs to be adjusted to consider specific aspects of the epidemic under analysis [25]. First, we predicted the SEIRV model dynamics without vaccination. Thereafter, the predictions were based on, evolution of infectious proportion under different levels of vaccination coverages (5%, 15%, 30%, 45%, 60%, 75%, 90%), SEIRV under different levels of vaccine efficacies (40%, 60%, 80%) with the current rate at different R_0_ (2,4,6) and SEIRV under combined coefficient (vaccine rate x vaccine efficacy) different levels of vaccination rates (1%, 2%, 5%), different vaccine efficacies (40%, 60%, 80%) and different R_0_ (2,4,6).

### Prediction based on the SEIRV model without vaccination

We observed how SEIR dynamics are affected without vaccination for COVID-19 at a specific time in the system’s evolution with different R_0_ values. When the R_0_ equals 2, 4 and 6, the susceptibility will be reduced around 150 days, 75 days, and 50 days respectively. Thus, the number of days to reach the peak of the infection curve will be 230 days, 105 days, and 74 days respectively. In addition, approximately 2.4 million individuals, 6.2 million individuals and 8.1 million individuals will be infected at the peak of the infection curves, respectively. Thus, around 0.97 million individuals in 225 days, 3 million individuals in 90 days and 8.1 million individuals in 74 days will be exposed at the peak of the exposed curves, respectively. Furthermore, the susceptible curve crosses with the recovered curve in 230 days with 10 million individuals, 95 days with 6 million individuals and 70 days with 4 million individuals, respectively. Moreover, the susceptible curves stabilize around 260 days with susceptible 4 million individuals, 115 days with susceptible 0.1 million individuals and 75 days with susceptible 0.05 million individuals, respectively. Furthermore, the recovered curves stabilize with 15 million around 300 days, 21.8 million around 125 days and 21.9 million individuals around 100 days, respectively (Fig 2 and S1 Table).

**Fig 2.**
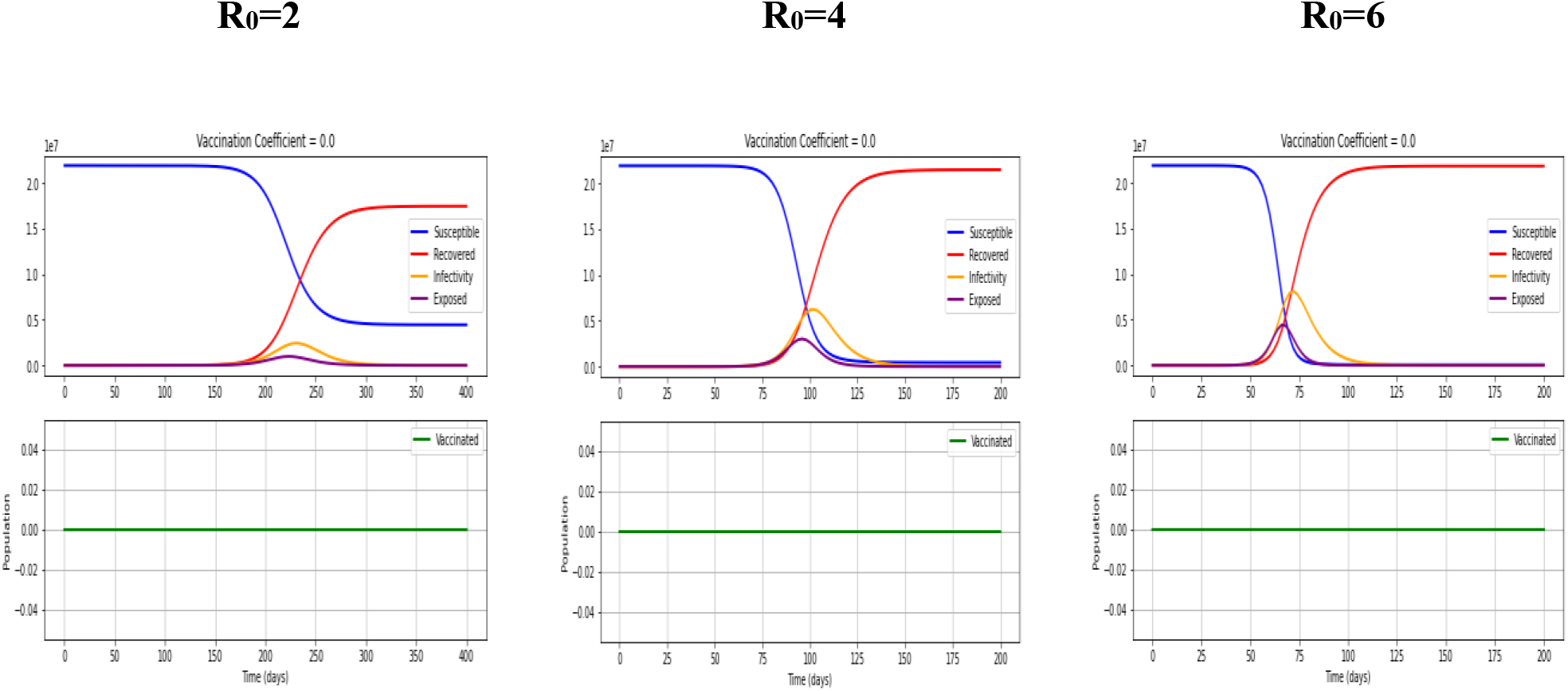
Evolution of infectious proportion without vaccination with different R_0_ values. Figure 2 is an ‘Evolution of infectious proportion without vaccination with different R_0_ values’.

**S1 Table. Evolution of Infectious and Exposed populations without vaccination**.

Supplementary Table 1 is an ‘Evolution of Infectious and Exposed populations without vaccination’.

### Evolution of infectious proportion under different levels of vaccination coverage

Using the SEIRV model, we predicted the proportion of the infected population for the different vaccination coverages taking into account the vaccination rate of the first 100 days in Sri Lanka (Per day vaccination rate=0.06%). According to the findings, if there are 5%, 15%, 30%, 45%, 60%, 75% and 90% vaccination coverages, the time of reaching the peak proportion of the infected individuals will be reduced by 40, 30, 25, 22, 21, 20 and 19 days respectively (Table 3). Moreover, after 45% vaccine coverage of the susceptible individuals, there will be a relatively slow reduction of peak reach for the proportion of the infected population. Therefore, at least 45% fully vaccine coverage will be adequate for reducing the infected population to control the outbreak since there is no prior immunity in the local community.

**Table 3.** Relationship between different vaccination coverages with the infected proportion and time of peak arrival.

| Percentage of vaccine coverage | Proportion infected out of<br>Susceptible (21.9 million) | Time of peak arrival |
| --- | --- | --- |
| 5% | 6.80% | Day 40 |
| 15% | 2.70% | Day 30 |
| 30% | 1.80% | Day 25 |
| 45% | 1.60% | Day 22 |
| 60% | 1.50% | Day 21 |
| 75% | 1.45% | Day 20 |
| 90% | 1.44% | Day 19 |

### Evolution of SEIRV under different vaccine efficacies and different R_0_ (2,4,6) with per day vaccination rate of 0.06% (First 100 days)

We observed how the dynamics of SEIR are affected by vaccination for COVID-19 at a specific time in the system’s evolution in the first 100 days after initiation of the vaccination campaign in Sri Lanka. As a result, vaccines with different efficacies as a response strategy applied to the COVID-19 epidemic was assumed at different R_0_ values.

### R_0_=2; Efficacy= 40%, 60%, 80% with per day vaccination rate 0.06%

When R_0_ is equal to 2, with 40%, 60% and 80% vaccine efficacies at the current rate of vaccination (0.06% per day), 80,256, 119,792 and 158,502 individuals can be vaccinated within 50 days, respectively. The susceptibility will be reduced to around 175 days, 160 days, and 150 days and the number of days to reach the peak of the infection curve will be 240 days, 245 days, and 250 days respectively. Moreover, 2.5 million individuals, 3 million individuals and 2.5 million individuals will be infected at the peak of the infection curves, respectively. Furthermore, 1.5 million individuals in 230 days, 0.5million individuals in 235 to 240 days and 2.5 million individuals in 245 days will be exposed at the peak of the exposed curve. Moreover, the susceptible curves stabilize with susceptible 5 million in 290 days, 4.5million in 280 days and 4 million individuals in 275 days correspondingly. In addition, the recovered curves stabilize with 16 million individuals, 17 million individuals and 18 million individuals, respectively, with 40%, 60% and 80% vaccine efficacies (Fig 3; S2 Table).

**Fig 3.**
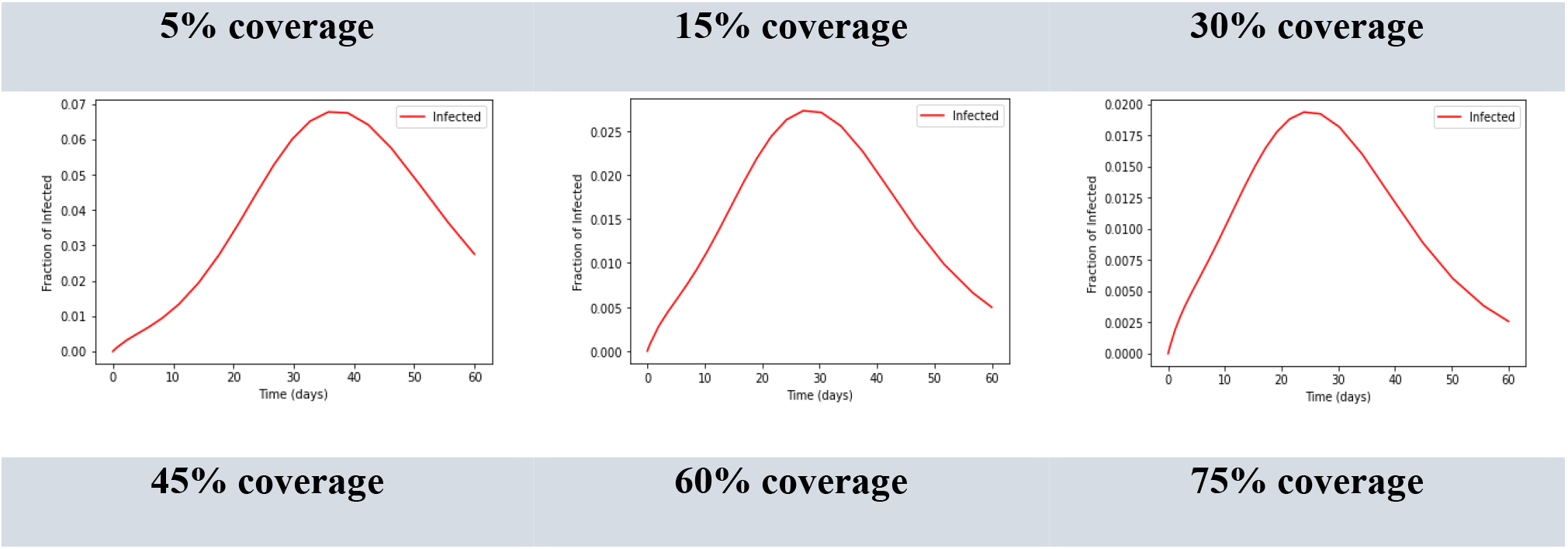
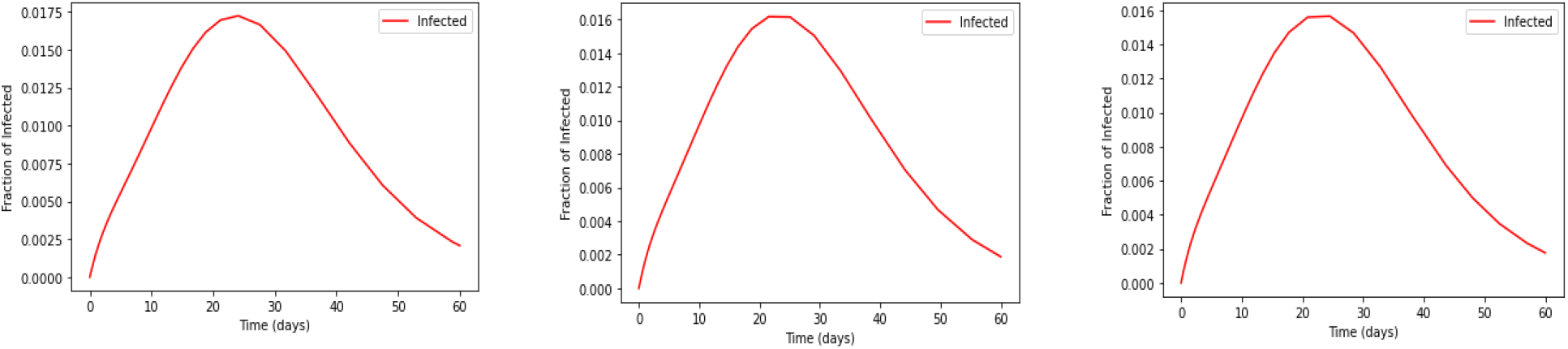
Prediction based on the SEIRV model considering the parameter R_0_ (R_0_=2,4,6) and Vaccine efficacy variation (Vaccine efficacy=40%,60%,80%)

Figure 3 is a prediction based on the SEIRV model considering the parameter R_0_ (R_0_=2,4,6) and Vaccine efficacy variation (Vaccine efficacy=40%,60%,80%)

**S2 Table. Evolution of SEIRV under different vaccine efficacies and R**_**0**_ **levels**

Supplementary Table 2 is an ‘Evolution of SEIRV under different vaccine efficacies and R_0_ levels’.

### R_0_=4; Efficacy= 40%, 60%, 80% with per day vaccination rate 0.06%

When R_0_ is equal to 4, with 40%, 60% and 80% vaccine efficacies at the current rate of vaccination (0.06% per day), 79,943, 119,144 and 157,852 individuals can be vaccinated in 50 days, and the susceptibility will be reduced around 65, 70 and 75 days respectively. Thus, the number of days to reach the peak of the infection curve will be 105, 110 and 115 days, and 5 million, 4.5 million and 4 million individuals will be infected at the peak of the infection curves, respectively. Furthermore, 2.5 million individuals in 90 days, 2 million individuals in 95 days and 1.5 million in 100 days will be exposed at the peak of the exposed curve, respectively. Correspondingly, the susceptible curve crosses with the recovered curve in 90 days with 5 million, in 95 days with 6 million and 6.5 million populations in 100 days. Moreover, the susceptible curves stabilize around 125 days with 0.5 million, 120 days with 0.4 million individuals and 115 days with susceptible 0.25 million individuals (Fig 3; S2 Table).

### R_0_=6; Efficacy= 40%, 60%, 80% with per day vaccination rate 0.06%

When R_0_ is equal to 6, with 40%, 60% and 80% vaccine efficacies at the current rate of vaccination (0.06% per day), 77,869, 116,289 and 154,364 individuals can be vaccinated in 40, 50 and 55 days respectively. The susceptibility will be reduced around 45, 50 and 55 days, respectively. Thus, the number of days to hold out the peak of the infection curve will be 70, 73 and 75 days, and 7.97 million, 7.89 million and 7.80 million individuals will be infected at the peak of the infection curves, respectively. Correspondingly, 4.29 million individuals in 65 days, 4.25 million individuals in 70 days and 4.22 million individuals in 75 days will be exposed at the peak of the exposed curve. Moreover, the susceptible curves stabilize around 75 days with 0.1 million individuals, in 80 days with 0.2 million individuals and 85 days with susceptible 0.3 million individuals, respectively (Fig 3; S2 Table).

### Evolution of SEIRV under different vaccine efficacies (40%, 60%, 80%) and different per day vaccination rates (1%, 2%, 5%)

The SEIRV predictions were performed for the different COVID-19 vaccine efficacies and the vaccination rates (per day) of 1%, 2% and 5% (Fig 4).

**Fig 4.**
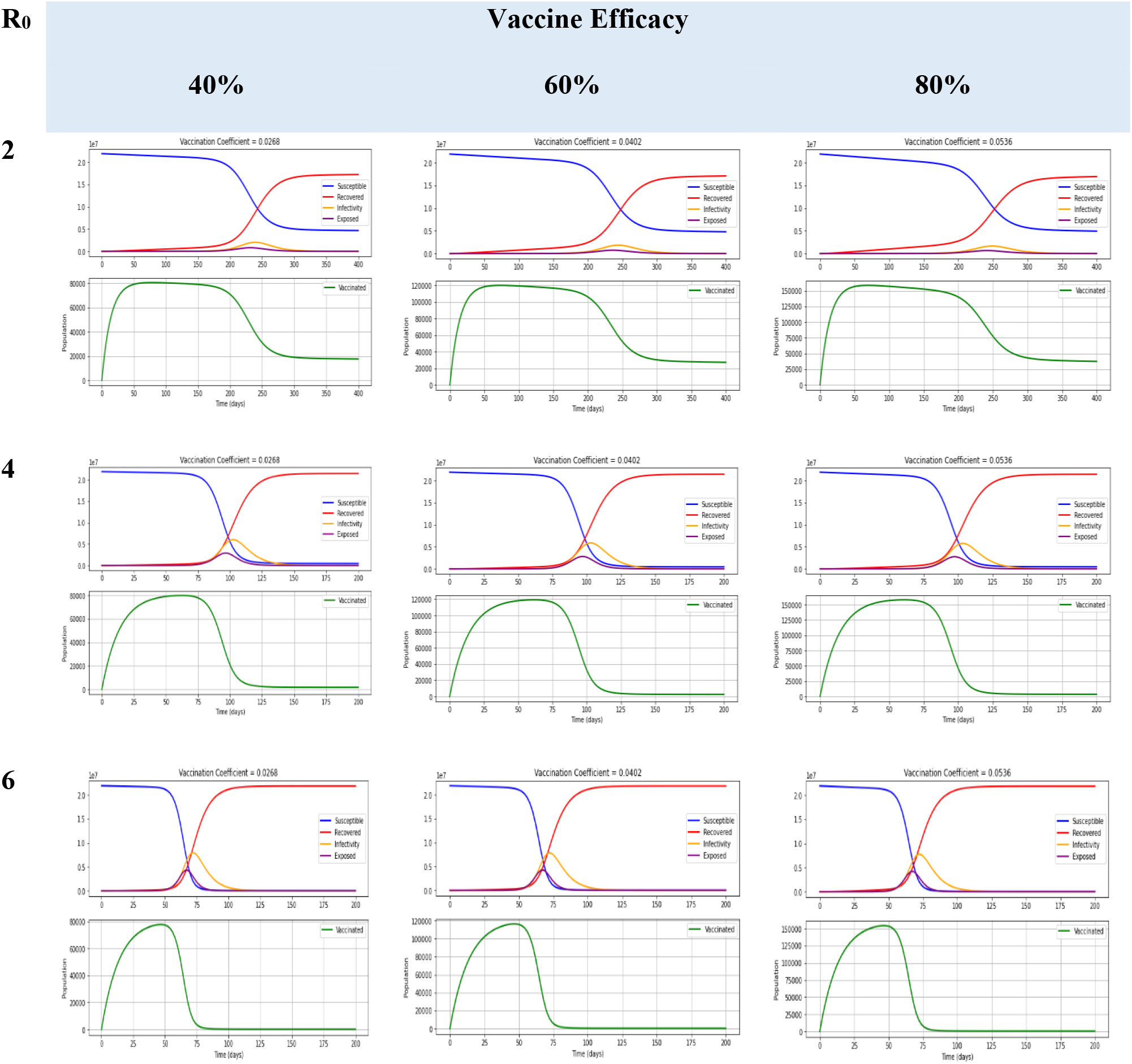
Evolution of SEIRV under different levels of vaccination rates (1%, 2%, 5%), vaccine efficacies (40%, 60%, 80%) and R_0_ of 2, 4 and 6.

Figure 4 is an ‘Evolution of SEIRV under different levels of vaccination rates (1%, 2%, 5%), vaccine efficacies (40%, 60%, 80%) and R_0_ of 2, 4 and 6’.

### R_0_=2; Efficacy= 40%; Vaccination rates of 1%, 2% and 5%

When R_0_ is 2 with vaccine efficacy of 40% and vaccination rates of 1%, 2% and 5%, 1.03 million, 1.86 million and 3.74 million individuals can be vaccinated in 35 days, 30 days, and 25 days respectively, and the susceptible curves cross the recovered curve in 180 days, 90 days, and 35 days respectively (Fig 4a; S3 Table).

**Fig 4a.**
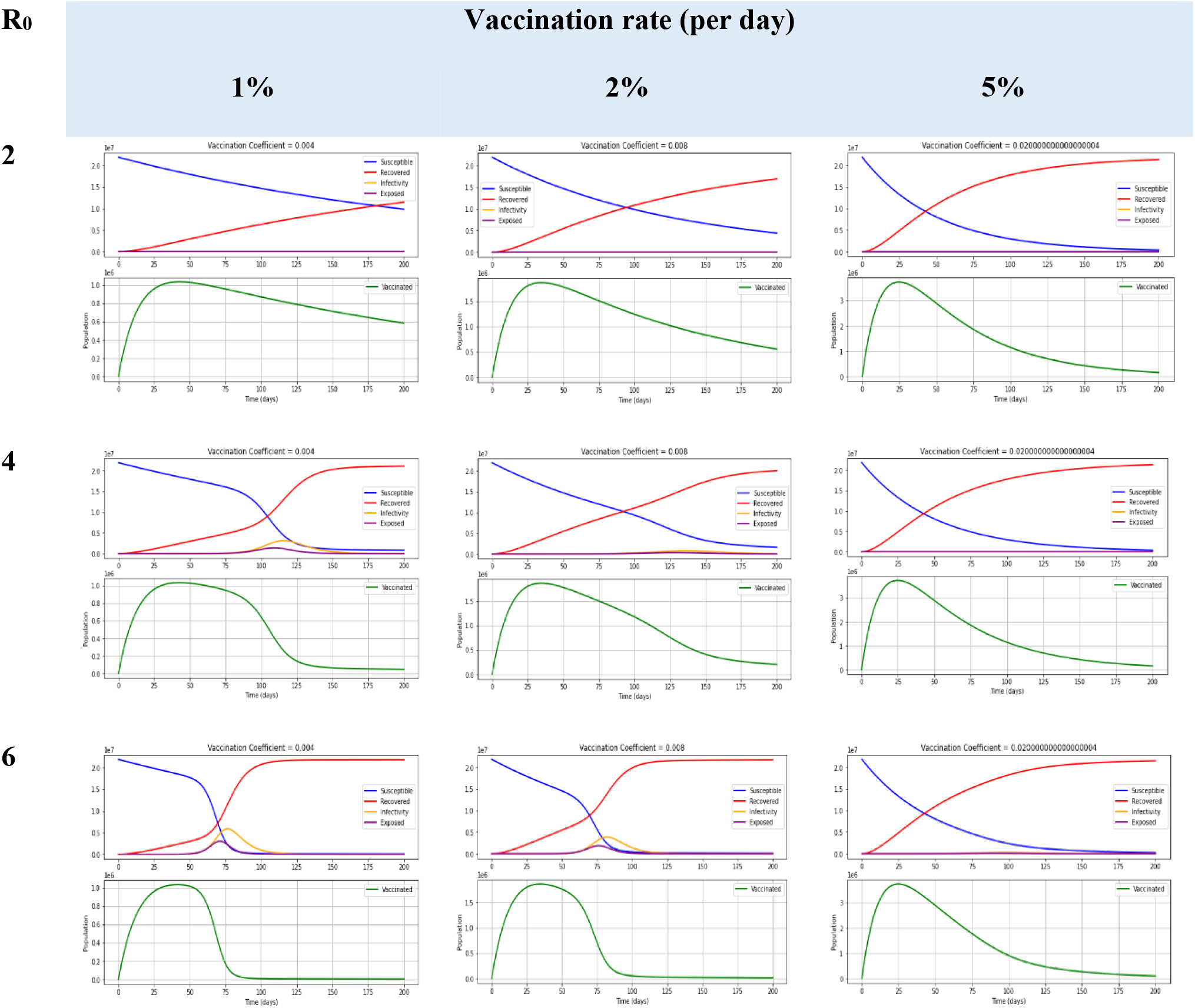
Evolution of SEIRV under different levels of vaccination rates (1%, 2%, 5%), R_0_ of 2, 4, 6 with 40% vaccine efficacy.

Figure 4a is an ‘Evolution of SEIRV under different levels of vaccination rates (1%, 2%, 5%), R_0_ of 2, 4, 6 with 40% vaccine efficacy’

**S3 Table. Evolution of SEIRV under different vaccination rates at R**_**0**_**=2,4,6 with the vaccine Efficacy of 40%, 60% and 80%**

Supplementary Table 3 is an ‘Evolution of SEIRV under different vaccination rates at R_0_=2,4,6 with the vaccine Efficacy of 40%, 60% and 80%’.

### R_0_=2; Efficacy= 60%; Vaccination rates of 1%, 2% and 5%

When R_0_ is equal to 2 with the efficacy of 60% and 1%, 2% and 5% vaccination rate (per day), 1.46 million, 2.57 million and 4.9 million individuals can be vaccinated in 30 days, 25 days, and 20 days respectively. Accordingly, the susceptible curve crosses with the recovered curve in 125 days, 65 days, and 30 days (Fig 4b; S3 Table).

**Fig 4b.**
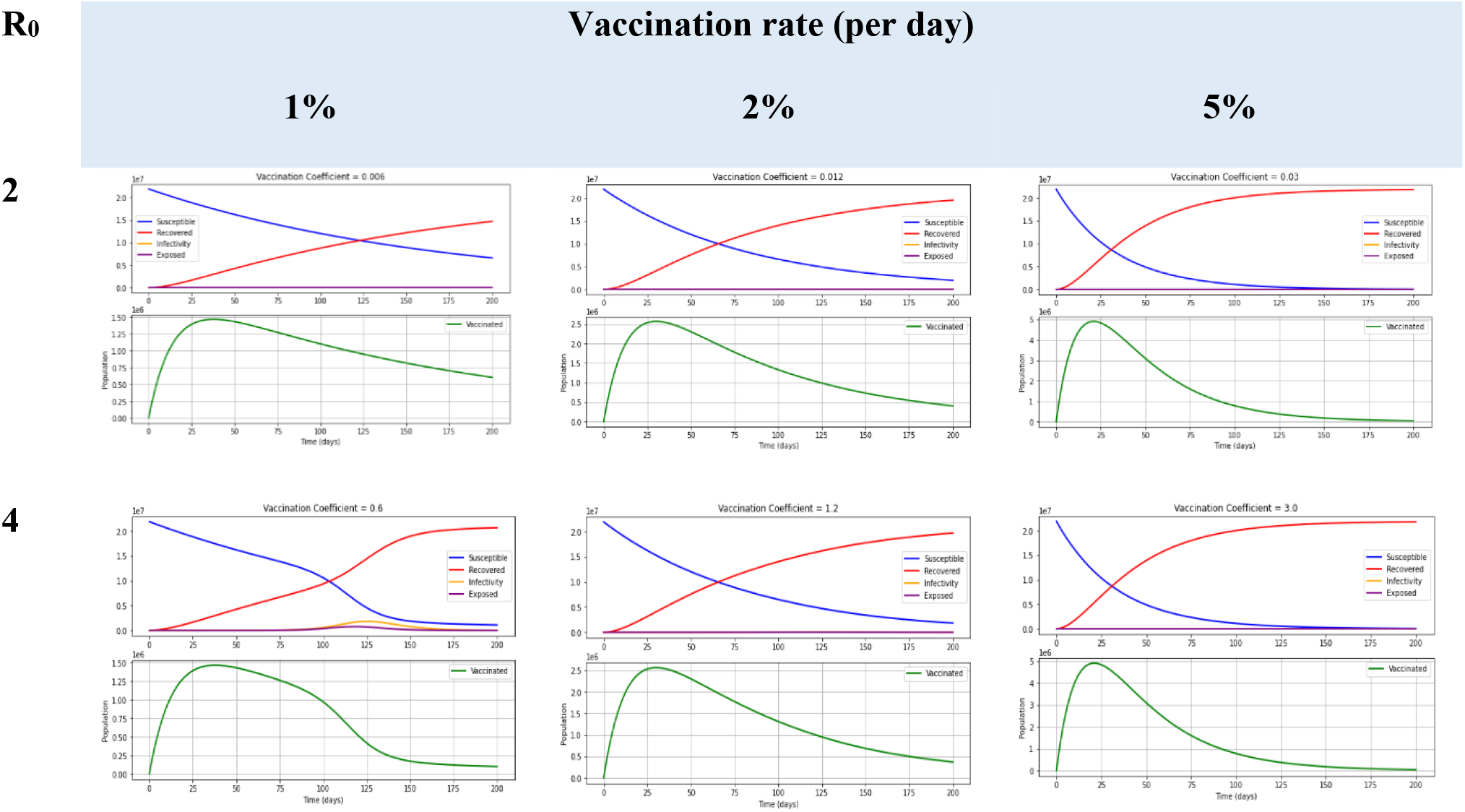
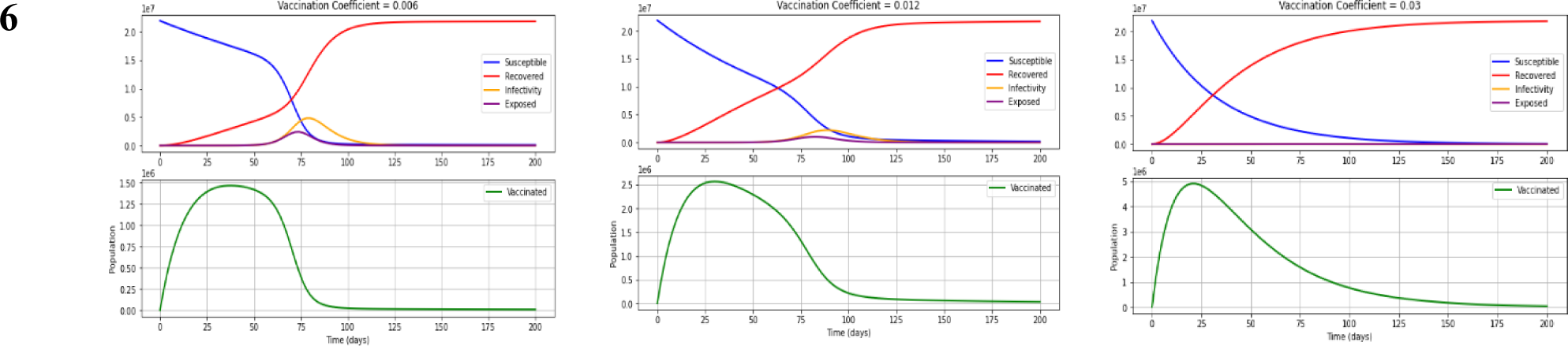
Evolution of SEIRV under different levels of vaccination rates (1%, 2%, 5%), R_0_ of 2, 4, 6 with 60% vaccine efficacy.

Figure 4b is an ‘Evolution of SEIRV under different levels of vaccination rates (1%, 2%, 5%), R_0_ of 2, 4, 6 with 60% vaccine efficacy’

### R_0_=2; Efficacy= 80%; Vaccination rates of 1%, 2% and 5%

When R_0_ is 2 with vaccine efficacy of 80% and vaccination rates of 1%, 2% and 5%, 1.86 million, 3.19 million and 5.87 million individuals can be vaccinated in 35 days, 25 days, and 15 days respectively. The susceptible curves cross with the recovered curve in 90 days, 50 days, and 25 days (Fig 4c; S3 Table).

**Fig 4c.**
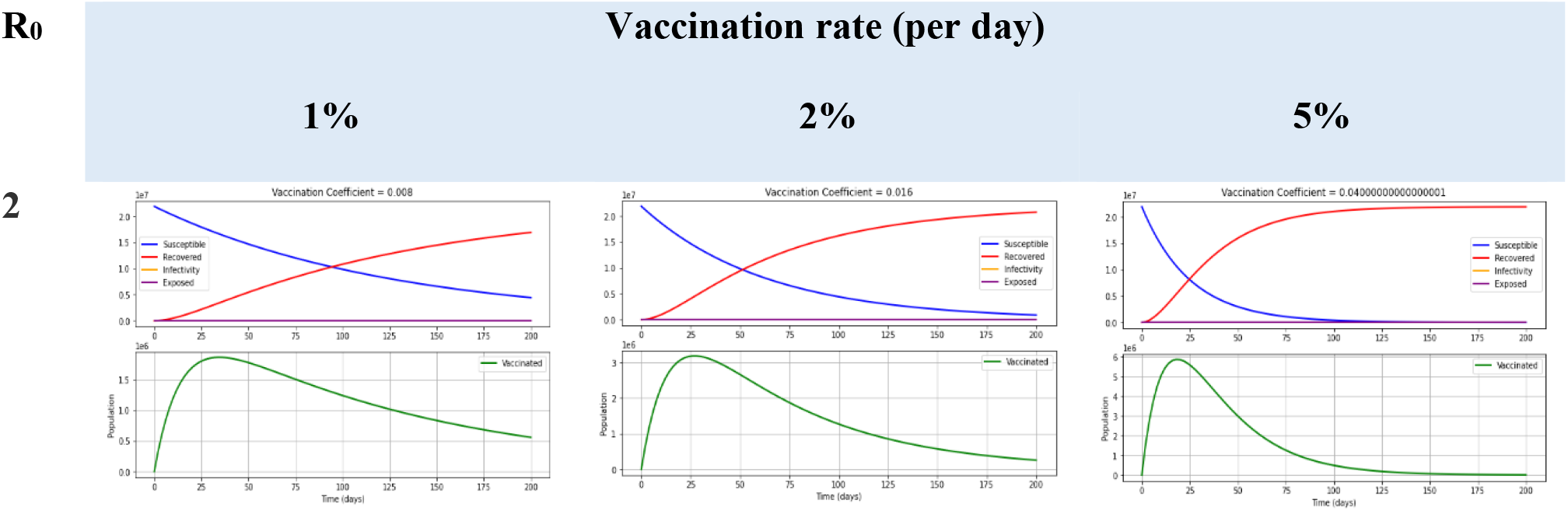
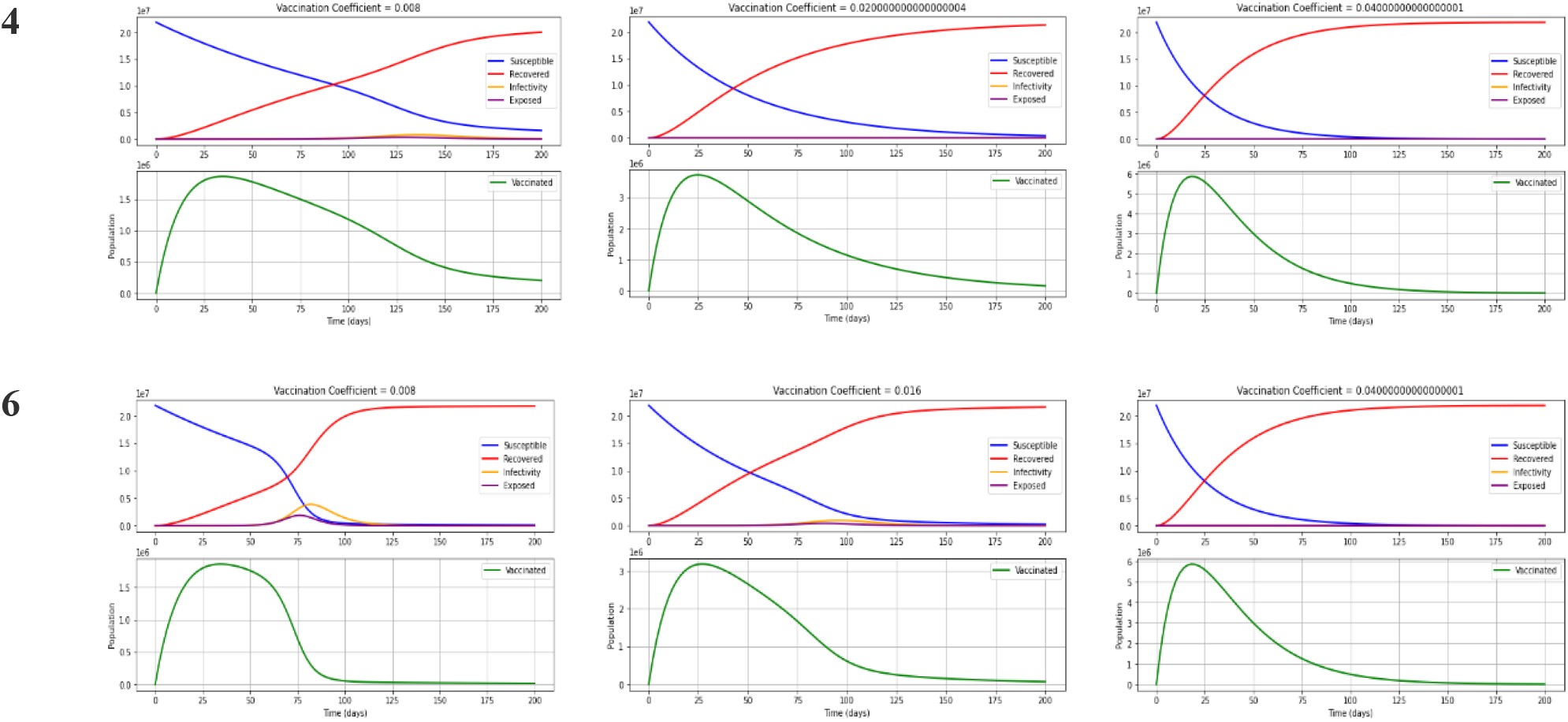
Evolution of SEIRV under different levels of vaccination rates (1%, 2%, 5%), R_0_ of 2, 4, 6 with 80% vaccine efficacy.

Figure 4c is an ‘Evolution of SEIRV under different levels of vaccination rates (1%, 2%, 5%), R_0_ of 2, 4, 6 with 80% vaccine efficacy’

### R_0_=4; Efficacy= 40%; Vaccination rates of 1%, 2% and 5%

When R_0_ is 4 with vaccine efficacy of 40% and vaccination rates of 1%, 2% and 5%, 1.03 million, 1.86 million and 3.74 million individuals can be vaccinated in 35 days, 35 days, and 25 days respectively. Moreover, 3.11 million, 0.81 million individuals and 1,478 individuals will be infected at the peak of the infection curves, and 1.36 million, 0.33 million, and 589 individuals will be exposed at the peak of the exposed curves, respectively. Furthermore, the susceptible curves cross the recovered curve in 105 days, 90 days, and 35 days, respectively (Fig 4a; S3 Table).

### R_0_=4; Efficacy= 60%; Vaccination rates of 1%, 2% and 5%

When R_0_ is equal to 4 with the efficacy of 60%, 1.47 million, 2.57 million and 4.9 million individuals can be vaccinated in 30 days, 28 days, and 20 days respectively. If there is a 1% vaccination rate, the number of days to reach the peak of the infection curve will be 125, and 1.8 million individuals will be infected at the peak of the infection curve, and 53,773 and 91 will be infected with 2% and 5% vaccination rates respectively. In addition, at the peak of the exposed curves, 0.76 million individuals, 21,041 individuals, 225 individuals will be exposed with 1%, 2% and 5% rates, respectively. Furthermore, the susceptible curves cross with the recovered curve in 100 days, 65 days, and 30 days with 10 million individuals (Fig 4b; S3 Table).

### R_0_=4; Efficacy= 80%; Vaccination rates of 1%, 2% and 5%

When the vaccine efficacy is 80% with the vaccination rates of 1%, 2%, and 5%, 1.86 million individuals, 3.19 million individuals and 5.87 million individuals can be vaccinated in 35 days, 25 days, and 20 days. Moreover, 0.81 million individuals in 135 days, 5,883, and 84 individuals will be infected at the peak of the infection curves, respectively. Furthermore, 0.33 million individuals in 130 days, 2,321 and 35 individuals will be exposed at the peak of the exposed curves. The susceptible curves cross with the recovered curve in 90 days, 40 days, and 25 days respectively, around 8 million individuals (Fig 4c; S3 Table).

### R_0_=6; Efficacy= 40%; Vaccination rates of 1%, 2% and 5%

When R_0_ is 6 with vaccine efficacy of 40% and vaccination rates of 1%, 2% and 5%, 1.03 million individuals, 1.86 million individuals and 3.74 million individuals can be vaccinated in 35 days, 30 days, and 25 days respectively. Moreover, 5.88 million individuals, 3.89 million individuals and 0.28 million individuals will be infected at the peak of the infection curves, and 3 million individuals, 1.88 million individuals and 0.11 million individuals will be exposed at the peak of the exposed curves. The susceptible curves cross with the recovered curve in 72 days with 7.5 million individuals, 70 days with 8 million individuals and 40 days with 10 million individuals, respectively (Fig 4a; S3 Table).

### R_0_=6; Efficacy= 60%; Vaccination rates of 1%, 2% and 5%

When R_0_ is equal to 6 with the efficacy of 60% and vaccination rates of 1%, 2%, and 5%, 1.47 million individuals, 2.57 million individuals and 4.9 million individuals can be vaccinated in 35 days, 30 days, 20 days, respectively. Moreover, 4.8 million individuals, 0.9 million individuals and 5,897 individuals will be infected at the peak of the infection curve, and 2.4 million individuals, 2.2 million individuals, and 14,420 individuals will be exposed at the peak of the exposed curves, respectively (Fig 4b; S3 Table).

### R_0_=6; Efficacy= 80%; Vaccination rates of 1%, 2% and 5%

When R_0_ is 6 with vaccine efficacy of 80% and vaccination rates of 1%, 2% and 5%, 1.86 million individuals, 3.19 million individuals and 5.87 million individuals can be vaccinated in 30 days, 25 days, and 20 days respectively. Moreover, 3.89 million individuals, 0.95 million individuals and 1,101 individuals will be infected at the peak of the infection curves, respectively. Besides, 1.88 million individuals, 0.4 million individuals and 2,644 individuals will be exposed at the peak of the exposed curves, respectively. The susceptible curves cross with the recovered curve in 70 days, 50 days, and 25 days (Fig 4c; S3 Table).

## Discussion

Predictive models have taken on a newfound importance in response to the spread of the COVID-19 pandemic and causative agent [26]. There are widespread public discussions on COVID-19 based on the features of epidemiological curves. For understanding the dynamics of the pandemic and assessing the effects of various intervention strategies, the epidemiological models and their graphical representations are valuable tools. However, the parameters may be affected by the inadequate explanations of these models’ representations, usefulness, and inherent limitations. Notably, accurate public interactions and communications are vital during any disastrous situation like the COVID-19 global pandemic. Moreover, explaining the current circumstances, actions, and intended outcomes is a timely need to gain the support and cooperation of the public and stakeholders to manage the critical situation, prevent the spreading of fake news, and minimize civil disobedience [27]. The compartmental models were invented during the late 1920s, which are the most commonly used models in epidemiology. Moreover, different approaches using agent-based simulations are still based on those basic models [25]. The SEIR model is frequently used to explain the COVID-19 pandemic, which is fundamental and a reasonably good fit for this disease [14]. Furthermore, the accuracy of the predictions of the epidemiological models depends critically on the quality of the parameters feed into the model. If the data quality is good, the model can precisely describe the situations. A fitting example would be when accurately estimating the case fatality rate, which requires all disease cases and the number of the dead [6]. However, during the COVID-19 pandemic, the number of deaths has often been highly inaccurate for many reasons, and the number of infected has also been incorrect. There can be undiagnosed cases during that period because of limited testing, which lead to inaccurate reporting [25,28]. The type of model is an extension of SEIR with the intervention of vaccination. The proposed model uses the predictors as in the parameter table (Table 2). The model was built based on the conceptual framework developed with the predictor selection. The model was internally validated using the parameters available in previous studies and situations related to New Delhi in India based on the literature. As with any modelling approach, our findings relate to the assumptions and inputs of the model, which lead to a major limitation. The assumptions with the greatest potential effect on our findings are the structural assumptions of a compartmental epidemiological model. Moreover, due to the static nature of the parameters, which does not reflect any internal or external change during the epidemic is a limitation of these models [8,28]. Furthermore, the predictive capability of the tool is highly dependent on several preliminary data for parameter estimation. This dependence may lead to data misinterpretation, especially considering the SIR model. Notably, an essential parameter in epidemic modelling is the ‘basic reproduction ratio (R_0_)’, which is the ‘expected or average number of individuals an infected person subsequently infects’. The size of the R_0_ can be varied since it is determined by averaging many cases. Moreover, R_0_ depends on the contagiousness of the pathogen and the number of contacts of an infected person [25]. When R_0_ and the rate of vaccination are constant, with increased vaccination efficacies, the vaccination coefficient is increased. It implies that the increase of vaccination coefficient would increase the vaccinated population while reducing the exposed and infectious population to reach its maximum with a low number of days. For instance, considering R_0_ equals 2 with 40% vaccine efficacy, the vaccination coefficient is 0.0268 (Fig 4). Thus, it needs to vaccinate 80,000 people within 50 days and continue to 200 days to reduce the exposed and infected populations. Moreover, with the constant R_0_, increase vaccination efficacies, and increased rate of vaccinations lead to increased vaccination coefficient. Therefore, it leads to an increased number of vaccinated populations from the susceptible population. As a result, the number of exposed, infectious populations are reduced and reach the equilibrium level at early days in the prediction graphs, which indicates that the COVID-19 crisis can be reduced if we increase the vaccine efficacy and the rate of vaccination. Under a lower level of vaccination coefficient, the equilibrium comes in later days of the prediction graphs while it increases the exposed and infectious populations. Therefore, we must give more attention to bring more efficient vaccines for COVID-19, and the rate of the vaccination campaigns need to be improved as a disaster mitigation strategy. In addition, the infection fatality ratio (IFR) of COVID-19 acts as a simple factor in the mortality effects of vaccination and does not alter the relative conclusions. The IFR estimates this proportion of deaths among all infected individuals. There are limited serological studies to calculate IFR accurately during outbreaks. In such situations, estimates need to be made with routinely available surveillance data, which generally consist of time series of cases and deaths reported in aggregate [29]. When considering the available data, it is almost similar to the study in China [21]. During the first 10 to 12 months, the mitigation policies reduced overall cumulative mortality without necessarily relying on the emergence of an effective vaccine. The model presented in this study revealed that the overall benefit of vaccination of a population is helping to suppress the epidemic curve by minimizing infected populations. However, the local benefits deteriorate with any additional prolongation of the vaccine development process and delaying availability. When transmission slows due to accumulated population immunity derived from infection, the benefit of vaccination needs to be assessed by conducting proper post-vaccination trials [30,31]. Moreover, the public health measures strongly influence the feasibility of vaccine trials during a COVID-19 epidemic. There is a natural tension between the goals of transmission control measures and field trial research because field trials rely on incident infection to demonstrate vaccine efficacy. The magnitude of potential benefits and risks of SARS-CoV-2 vaccine trials, potential benefits of post-trial vaccination campaigns and the marginal risks are influenced by the public health measures, impending disasters, and other epidemic conditions. Although high background risk of infection results in reduced marginal risks to participants, a substantially greater public health benefit results in low background risk during the period before vaccine availability [31,32]. The predictors of the SEIRV compartmental model have been analyzed using Ordinary Differential Equations (ODEs). The model predicts the plausible parameters with the robust estimation within the limitations. One of the significant limitations of the model is that it does not include the natural death and birth rates assuming those are constant [28,31]. Internal and external validation of the model is vital for the robust prediction of the ODEs in the model. Thus, the models were applied in the series of equations to get the equilibrium in the SEIRV model. Then, the simulation of the validated model was performed to obtain the policy scenarios of the proposed model. Usually, the SEIR model consists of initial parameters, which predicts as those are applied to the model precautionary [30,32]. Initially, the model comprises one exposed individual, and the rest of the population is considered a susceptible population [33,34]. Therefore, the predictors were handled with care in the model to avoid overestimation or underestimation.

## Implications

The prediction models will lead to policy relevance despite the significant uncertainty associated with real-time forecasting in complex systems with timely predictions and steadfast reports. Furthermore, new policy discussions need to occur whenever the best available options such as vaccination and knowledge about the epidemiology changes. Thus, the proposed model can serve as a tool for health authorities for planning and policymaking to control the pandemic by cost-effectively implementing appropriate vaccination campaigns.

## Conclusion

A computational model for predicting the spread of COVID-19 by dynamic SEIRV model has been proposed in the study. At least 45% vaccine coverage is required for reducing the infected population as a measure of disaster mitigation in Sri Lanka. Theoretically, R_0_ is varied [35], which estimates the speed at which a disease can spread in a population. Unfortunately, we didn’t estimate the reproductive ratios from data in a particular population which is useful for that population. However, the parameters such as vaccination efficacy and vaccination rate can be adjusted according to the implementation of preventive strategies. Therefore, the R_0_ is increased in the SEIRV model along with the increase of the vaccination efficacy. Moreover, the population to be vaccinated is also decreased for respective R_0_. Furthermore, when R_0_ is increased in the SEIRV model along with the increase of both vaccination efficacy and vaccination rate, the population to be vaccinated is also declined. Hence, the vaccination offers the greatest benefits to the local population by reducing the time of peak arrival and infected population.

## Data Availability

We extracted publicly available data with permission from the official website of the Health Promotion Bureau of the Ministry of Health, Sri Lanka and the data was cross-checked with the daily update on the website of the Epidemiology Unit, Ministry of Health, Sri Lanka

## Supporting Information

### Model Validation

Since the situation of New Delhi, India was similar to the Sri Lankan situation of the COVID-19 during the first quarter of 2021, we used the same data for validating the present SEIRV model. In addition, the time frame of the New Delhi situation is also similar to the Sri Lankan conditions (a parallel time frame). Accordingly, the validation component has supplemented the model that we have used in the analysis. Furthermore, we have considered R_0_ of 2, 4, 6 in this analysis.

**Table 1.** Model parameters.

| Parameter | Value (New Delhi) | Value (Sri Lanka) |
| --- | --- | --- |
| Incubation period | 5.74 | 5.5 |
| Hospitalization period | 21 | 22 |
| Population | 30,291,000 | 21,919,000 |
| $R_0$ | 2, 4, 6 (Assumed) | 2,4,6 (Assumed) |
| Vaccination rate 2% | 2 % | 1%, 2%, 5% |
| Vaccine efficacy | 60% (Assumed: AstraZeneca) | 40%, 60%, 80% |
| Social distancing and other policies | 0 | 0 |

Source: CDC (www.cdc.gov)

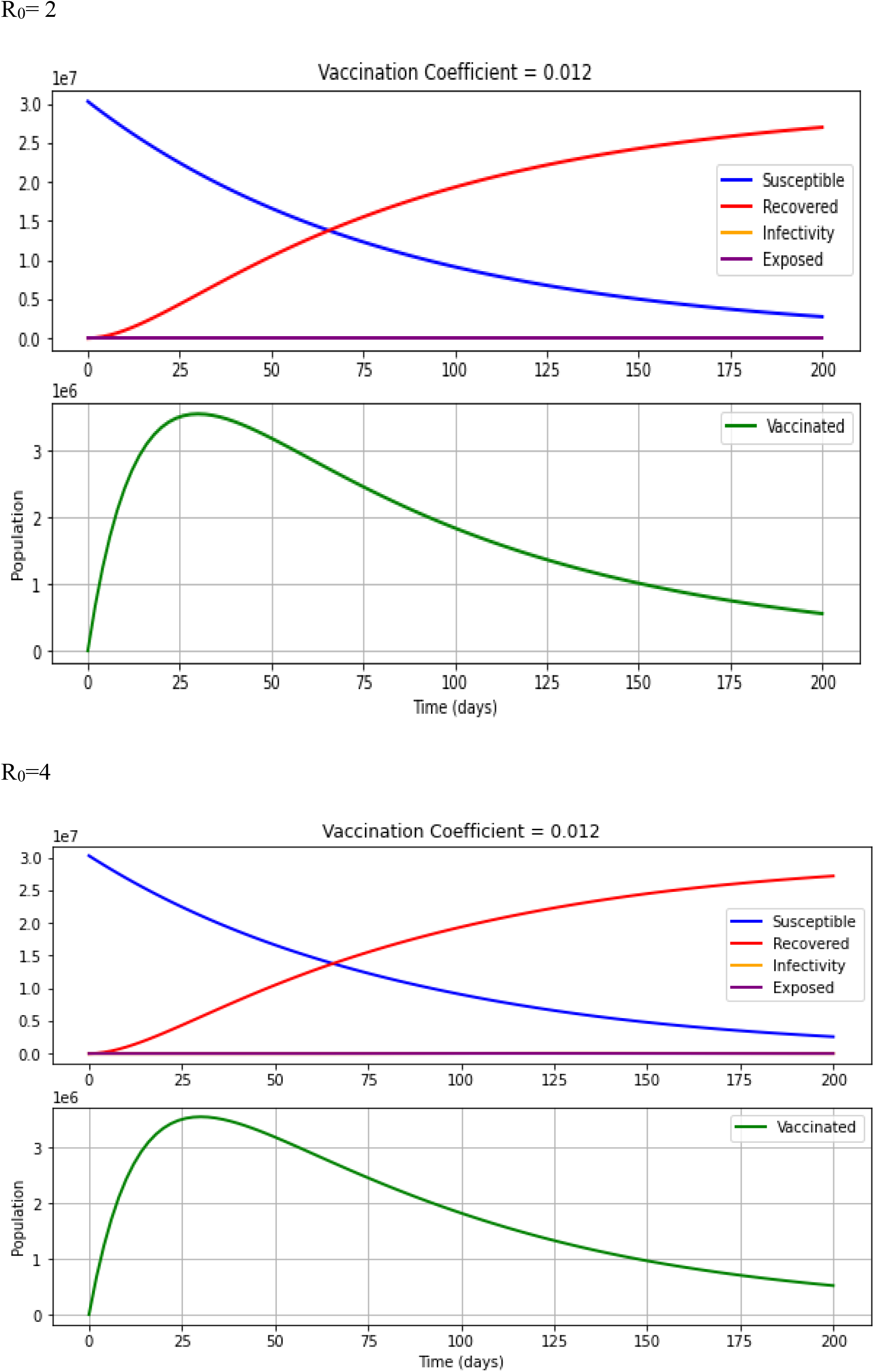
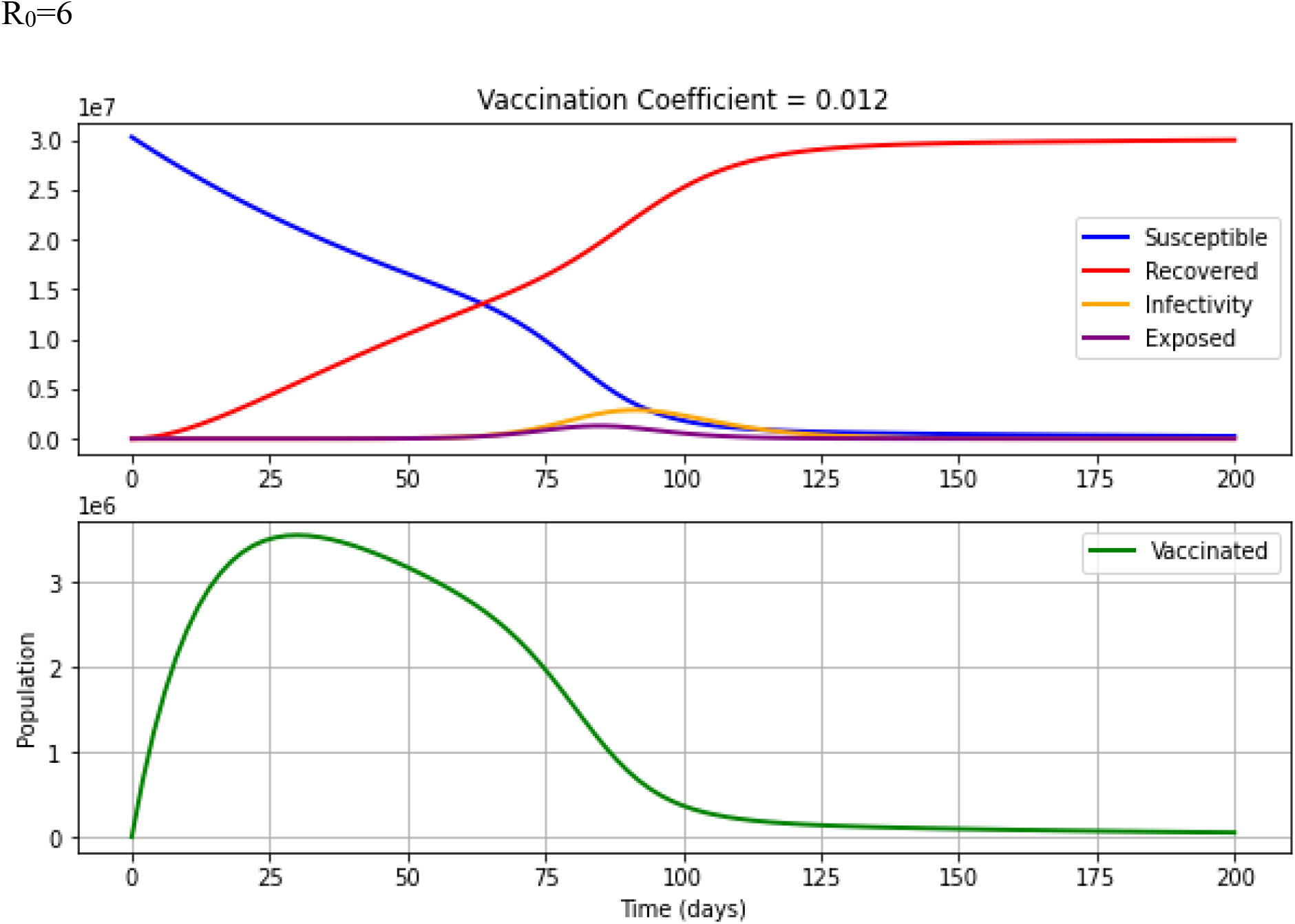

**Table 2.** SEIR model validation and simulation for New Delhi in India (Population)

| $R_0$ | Infection | Exposure | Vaccinated |
| --- | --- | --- | --- |
| $R_0=2$ | 11 | 5 | 3,549,725 |
| $R_0=4$ | 54,993 | 21,503 | 3,549,705 |
| $R_0=6$ | 2,872,463 | 1,286,209 | 3,549,523 |

## Author contributions

Conceptualization and methodology: MSDW, RMNUR, SPJ, IG, PCW; Software; SPJ; Formal analysis; SPJ and RMNUR; Original draft preparation; RMNUR; Writing; RMNUR, BMWIG, WMPCW; Review & editing; MSDW, RMNUR, SPJ, TKT, YA, SB.

